# A computational approach to identify interfering medications on urine drug screening assays without data from confirmatory testing

**DOI:** 10.1101/2020.04.03.20052621

**Authors:** Nadia Ayala-Lopez, Layla Aref, Jennifer M. Colby, Jacob J. Hughey

**Affiliations:** Department of Pathology, Microbiology, and Immunology, Vanderbilt University Medical Center, Nashville, Tennessee, USA; Department of Biomedical Informatics, Vanderbilt University Medical Center, Nashville, Tennessee, USA

**Keywords:** urine drug screening, electronic health records, statistical analysis

## Abstract

**Background:** Urine drug screening (UDS) assays can rapidly and sensitively detect drugs of abuse, but can also produce spurious results due to interfering substances. We previously developed an approach to identify interfering medications using electronic health record (EHR) data, but the approach was limited to UDS assays for which presumptive positives were confirmed using more specific methods. Here we adapted the approach to search for medications that cause false positives on UDS assays lacking confirmation data.

**Methods:** From our institution’s EHR data, we used our previous dataset of 698,651 UDS and confirmation results. We also collected 211,108 UDS results for acetaminophen, ethanol, and salicylates. Both datasets included individuals’ prior medication exposures. We hypothesized that the odds of a presumptive positive would increase following exposure to an interfering ingredient independently of exposure to the assay’s target drug(s). For a given assay-ingredient pair, we quantified potential interference as an odds ratio from logistic regression. We evaluated interference of selected compounds in spiking experiments.

**Results:** Compared to the approach requiring confirmation data, our adapted approach showed only modestly diminished ability to detect interfering ingredients. Applying our approach to the new data, three ingredients had a higher odds ratio on the acetaminophen assay than acetaminophen itself did: levodopa, carbidopa, and entacapone. The first two, as well as related compounds methyldopa and alpha-methyldopamine, produced presumptive positives at < 40 μg/mL.

**Conclusions:** Our approach can reveal interfering medications using EHR data from institutions at which UDS results are not routinely confirmed.

## Introduction

Urine drug screening (UDS) assays play a role in various clinical contexts, from the emergency department to outpatient rehabilitation and treatment centers. Because UDS assays prioritize sensitivity over specificity, positive UDS results can occur due to interference by non-targeted substances, e.g., other medications [1]. For this reason, positive UDS results are considered presumptive until confirmed by a more specific technique such as mass spectrometry. Results of confirmatory testing, however, are often not available until several days later, and many hospitals do not confirm presumptive positives at all. Better knowledge of the substances that cause false positives would help laboratorians and physicians who rely on UDS results to guide patient care.

Recently we developed and validated an approach, based on statistical analysis of electronic health record (EHR) data, to identify interfering medications that cause false positive UDS results [2]. In this initial work we relied on confirmation data to determine whether each presumptive positive was a true positive or a false positive. For the hospitals where presumptive positives are not automatically confirmed, however, our approach is not applicable.

In this study we extended our approach to identify medications capable of causing false positives on UDS assays that lack confirmation data. At our institution, this includes the assays for acetaminophen, ethanol, and salicylates. We applied our approach to approximately 5 years’ worth of UDS results and co-occurring medication exposures documented in the EHR, then validated the top hits experimentally.

## Materials and Methods

Code and summary-level data for this study are available at https://doi.org/10.6084/m9.figshare.12067233. The Vanderbilt Institutional Review Board reviewed and approved this study as non-human subjects research (IRB# 081418 and 190165).

### Extraction of UDS results and drug exposures from electronic health record data

EHR data came from the Synthetic Derivative, a collection of deidentified clinical data from Vanderbilt University Medical Center (VUMC) [3]. In addition to using the dataset from our previous study [2], we made a new dataset consisting of all UDS results for the currently used assays for acetaminophen, ethanol, and salicylates. The acetaminophen (Sekure Chemistry) and salicylates (Abbott Multigent) assays were validated as laboratory-developed tests, as they are not FDA-cleared for use in urine. The ethanol assay (Abbott Multigent) was FDA-cleared for use in urine. All testing was performed on an Abbott Architect c16000 automated chemistry analyzer.

For each person in the new dataset, we identified drug exposures documented between 1 and 30 days prior to each UDS result. We excluded UDS results that occurred less than 30 days after the person’s first ever visit at VUMC, since we would lack a prior 30 days of documented drug exposures. Documented drug exposures are available as structured data in the Synthetic Derivative and come primarily from medication lists. We mapped each drug to its active ingredient(s) using RxNorm [4]. As described previously, having a documented exposure within 30 days is only a proxy for being exposed at the time of the UDS [2].

### Statistical analysis of drug exposures and UDS results

We quantified associations between drug exposures and UDS results using Firth’s logistic regression [5,6]. Given the coefficients and standard errors from the logistic regression fits (where each coefficient corresponded to a log odds ratio), we then used an Empirical Bayes approach called adaptive shrinkage to estimate the posterior mean of the log odds ratio and the corresponding 95% credible interval for each assay-ingredient pair [7]. The latter is analogous to a confidence interval, but for Bayesian statistics.

For the re-analysis of our previous dataset, we fit two types of logistic regression models. In model 1, the dependent variable corresponded to the UDS result (negative or false positive) and the independent variable corresponded to presence or absence of prior exposure to the ingredient. In model 2, the dependent variable corresponded to the UDS result (negative or presumptive positive) and the independent variables corresponded to (i) presence or absence of prior exposure to the ingredient and (ii) presence or absence of prior exposure to the assay’s target drug(s) (if not the same as the ingredient of interest). For consistency with our previous study, we only fit a model for an assay-ingredient pair if exposure to the ingredient preceded a false positive (model 1) or presumptive positive (model 2) in at least five individuals.

For the analysis of our new dataset, we fit model 2 for assay-ingredient pairs for which exposure to the ingredient preceded a UDS result in at least 20 individuals. For the ingredients most strongly associated with presumptive positive results on the acetaminophen assay, we calculated co-exposure frequencies as the percentage of exposures to one ingredient for which the person was also exposed to a second ingredient.

### Experimental validation of interference

For each selected compound, we spiked a reference standard into drug-free urine at various concentrations and testing the spiked urine samples in singlicate on an Abbott Architect c16000 chemistry analyzer. We used linear interpolation to estimate the concentration of the test compound at which the assay registered a concentration equal to the cutoff.

We purchased reference standards from Tocris Bioscience (Bristol, UK). We prepared stock solutions of each standard in DMSO (carbidopa and entacapone), or in saline and HCl (levodopa). We spiked the urine samples using a fixed volume of 20% spiking solution, made of a combination of diluent and stock solution, including one sample per compound with only diluent to serve as a negative control. In most cases, we tested the maximum technically feasible concentration for a compound, given the limits of solubility, the concentration of the reference material, and the fixed 20% spiking volume.

## Results

We started with the dataset from our previous study [2], which included results from urine drug screens and confirmations for ten classes of target drugs, as well as each person’s prior documented drug exposures in terms of active ingredients (Supplemental Table 1). Without the confirmation results, one cannot know whether a given presumptive positive UDS result was a true positive or a false positive. We hypothesized, however, that the odds of a presumptive positive would increase following exposure to an interfering ingredient independently of exposure to the assay’s target drug(s).

We therefore used logistic regression followed by a technique called adaptive shrinkage [7] to quantify two types of associations: (1) between ingredient exposures and false positive UDS results, which used the confirmation as in our previous approach—yielding an odds ratio OR_FP_—and (2) between ingredient exposures and presumptive positive UDS results, adjusted for exposure to assay targets—yielding an odds ratio OR_P_ (Tables S2 and S3).

We compared OR_FP_ and OR_P_ for assay targets, previously known interferents, and “new” interferents that we discovered in our previous study (Figure 1). Consistent with our hypothesis, most interfering ingredients with a high OR_FP_ also had a high OR_P_ (Figure 1A-B). In addition, ranking by OR_P_ captured only moderately fewer interfering ingredients than ranking by OR_FP_ (Figure 1C). Thus, our approach can detect ingredients that may cause false positive UDS results, even if confirmation data are unavailable.

**Figure 1.**
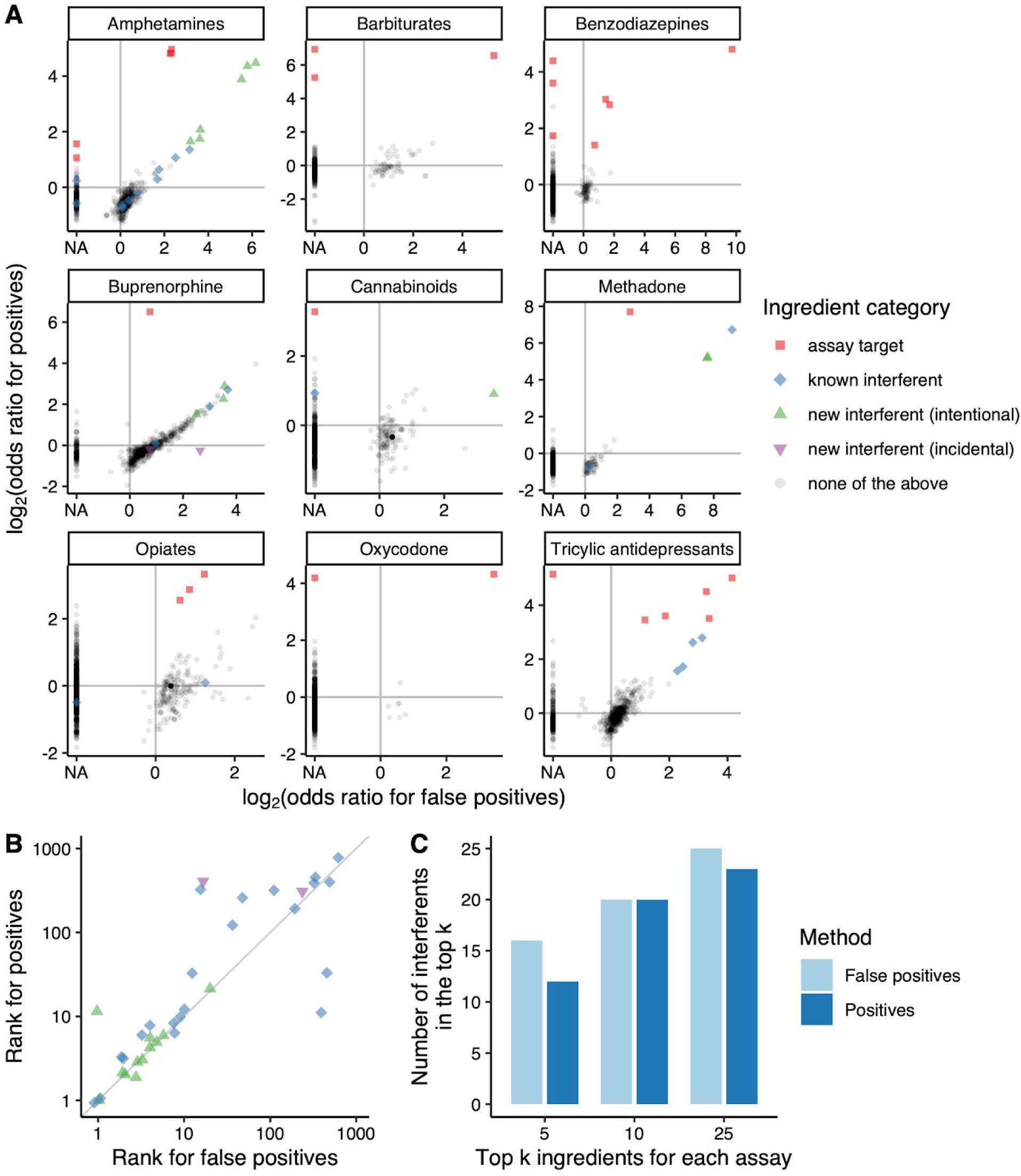
Comparison of calculating potential interference with and without confirmation data. **(A)** Each plot corresponds to a UDS assay, and each point corresponds to an ingredient. A log_2_(odds ratio for false positives) of NA indicates that the association was not tested because < 5 individuals had a false positive UDS result preceded by exposure to the given ingredient. The assay for cocaine metabolite is not shown, as it had no false positives. New interferent (intentional) indicates an ingredient we validated in our previous study based on its high odds ratios for false positives on the given assay. New interferent (incidental) indicates an ingredient we validated on a different assay than the one for which it had a high odds ratio. **(B)** Ranks of interfering ingredients on all assays based on odds ratio for false positives and odds ratio for presumptive positives. Ranks were calculated on a per-assay basis and excluded assay targets. Points are slightly jittered to avoid overlap. **(C)** Number of interfering ingredients across all assays with an odds ratio in the top k for each assay (where k = 5, 10, or 25), excluding assay targets.

We next assembled a dataset of UDS results for acetaminophen, ethanol, and salicylates, three assays for which presumptive positive results at our institution are not confirmed. The dataset included 211,108 results from 39,658 individuals (Table 1), along with each person’s documented drug exposures occurring between 1 and 30 days prior. Each UDS result was preceded by exposure to a median of 2 ingredients and a mean of 7.7 ingredients.

**Table 1.** Characteristics of newly analyzed urine drug screening assays.

| Target drug | Format | Manufacturer / Brand | Cutoff (µg/mL) | Number of UDS results |  |
| --- | --- | --- | --- | --- | --- |
|  |  |  |  | Negative | Presumptive positive |
| acetaminophen | Enzymatic (acyl amidohydrolase) / colorimetric | Sekure Chemistry / L3K Assay | 3 | 54,220 | 16,180 |
| ethanol | Enzymatic (alcohol dehydrogenase) / colorimetric | Abbott MULTIGENT | 100 | 65,012 | 5,473 |
| salicylates | Enzymatic (salicylate hydroxylase) / colorimetric | Abbott MULTIGENT | 5000 | 62,692 | 7,531 |

Using this new dataset, we calculated OR_P_ for 2,563 assay-ingredient pairs. Further supporting our approach’s validity, acetaminophen was the fourth-ranked ingredient on the acetaminophen assay and aspirin was the top-ranked ingredient on the salicylates assay (Figure 2). Ethanol as an ingredient had only 16 exposures in our dataset and showed no clear association with results on the ethanol assay (Supplemental Table 4).

**Figure 2.**
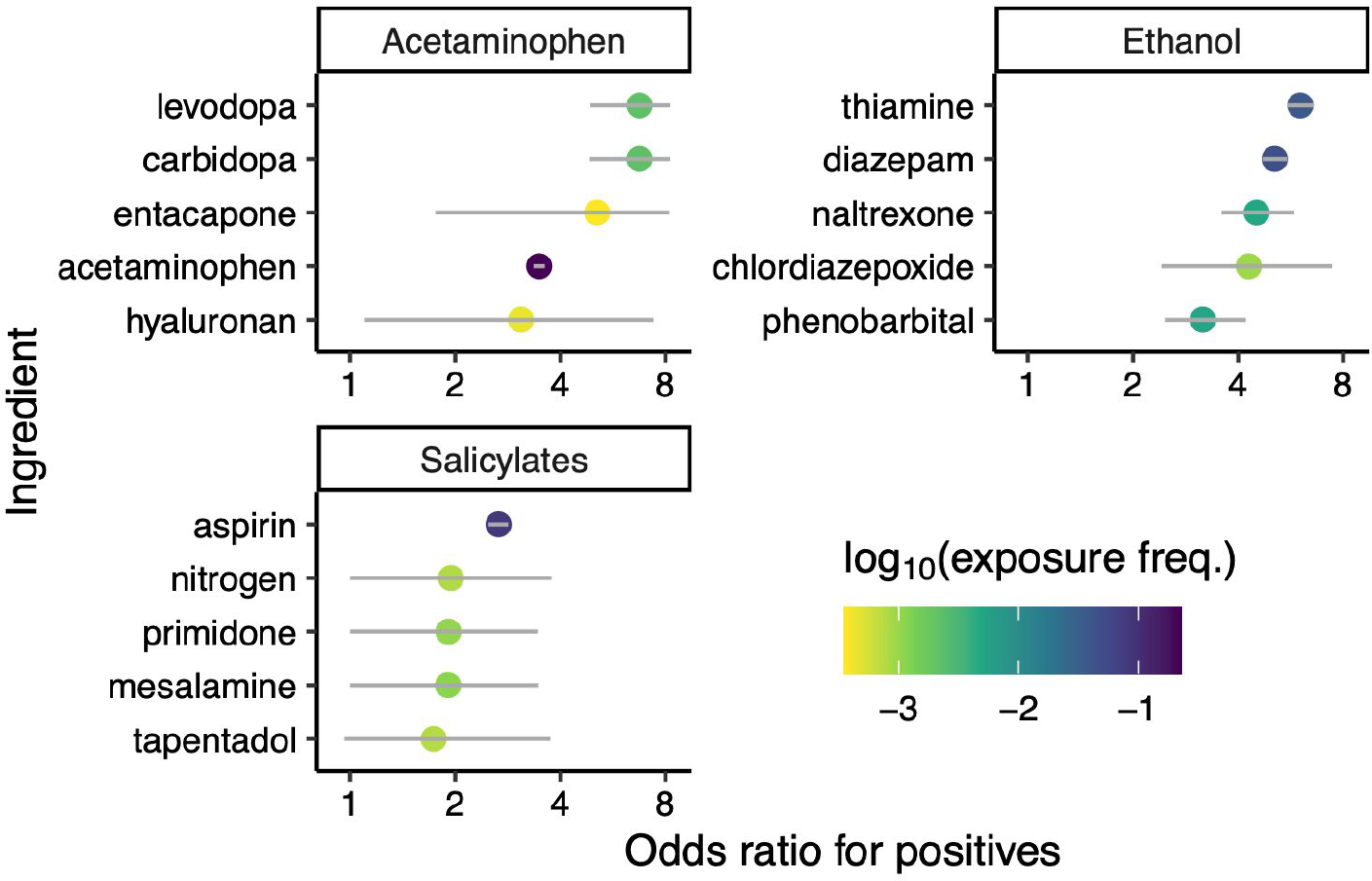
Top-ranked ingredients associated with presumptive positive UDS results for acetaminophen, ethanol, and salicylates. Gray lines over each point indicate 95% credible intervals. Exposure frequency corresponds to the fraction of UDS results preceded by exposure to the given ingredient. All tested associations for the three assays are in Supplemental Table 4.

The three ingredients that had a higher OR_P_ than acetaminophen on the acetaminophen assay were levodopa, carbidopa, and entacapone (Figure 2). These associations were unlikely to be due solely to co-exposure with acetaminophen, as each logistic regression model already accounted for exposure to the respective assay’s target drug. Co-exposure analysis indicated that the associations of levodopa and carbidopa were indistinguishable, because the two ingredients were almost always given together (Figure 3). Subsequent logistic regression also suggested that entacapone’s association with presumptive positive UDS results could be explained by co-exposure with levodopa and/or carbidopa (Supplemental Table 5).

**Figure 3.**
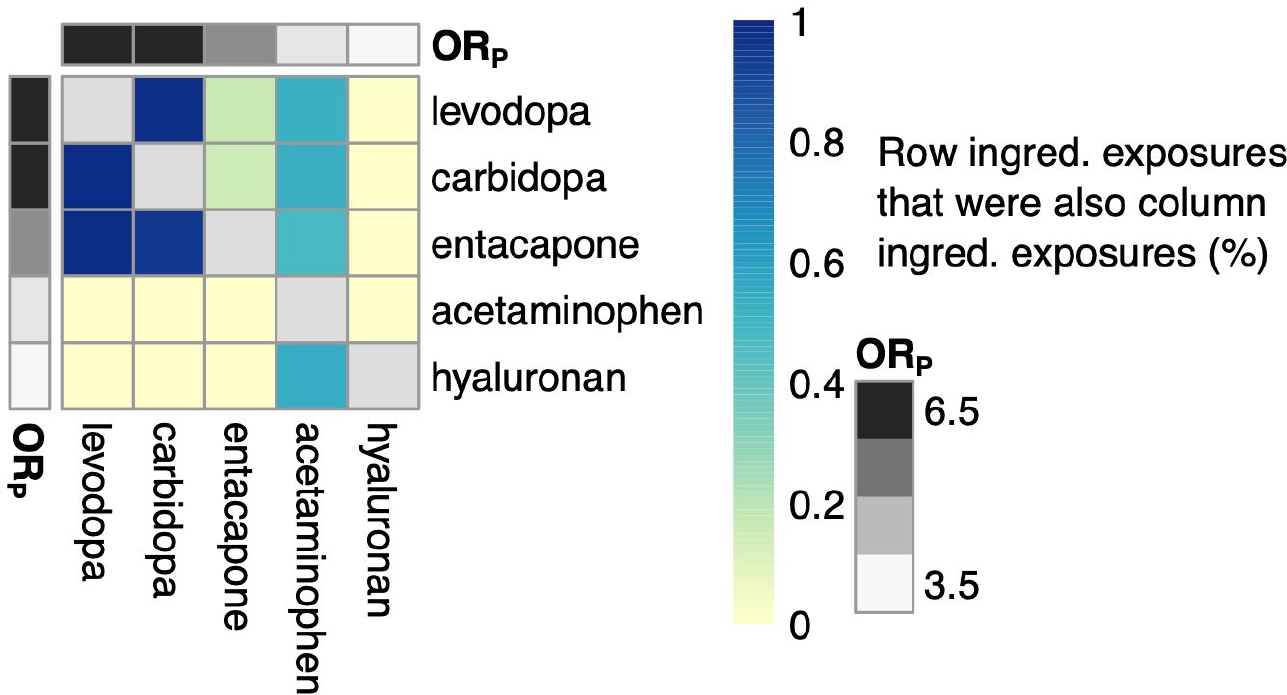
Co-exposure frequencies for the five ingredients most strongly associated with presumptive positive UDS results on the acetaminophen assay.

We evaluated the interference of each of these three compounds experimentally (Figure 4). Consistent with our analysis of the EHR data, both levodopa and carbidopa interfered strongly on the acetaminophen assay, each producing a presumptive positive (corresponding to an acetaminophen concentration of 3 μg/mL) at less than 40 μg/mL. Entacapone, on the other hand, produced a presumptive positive at 400 μg/mL.

**Figure 4.**
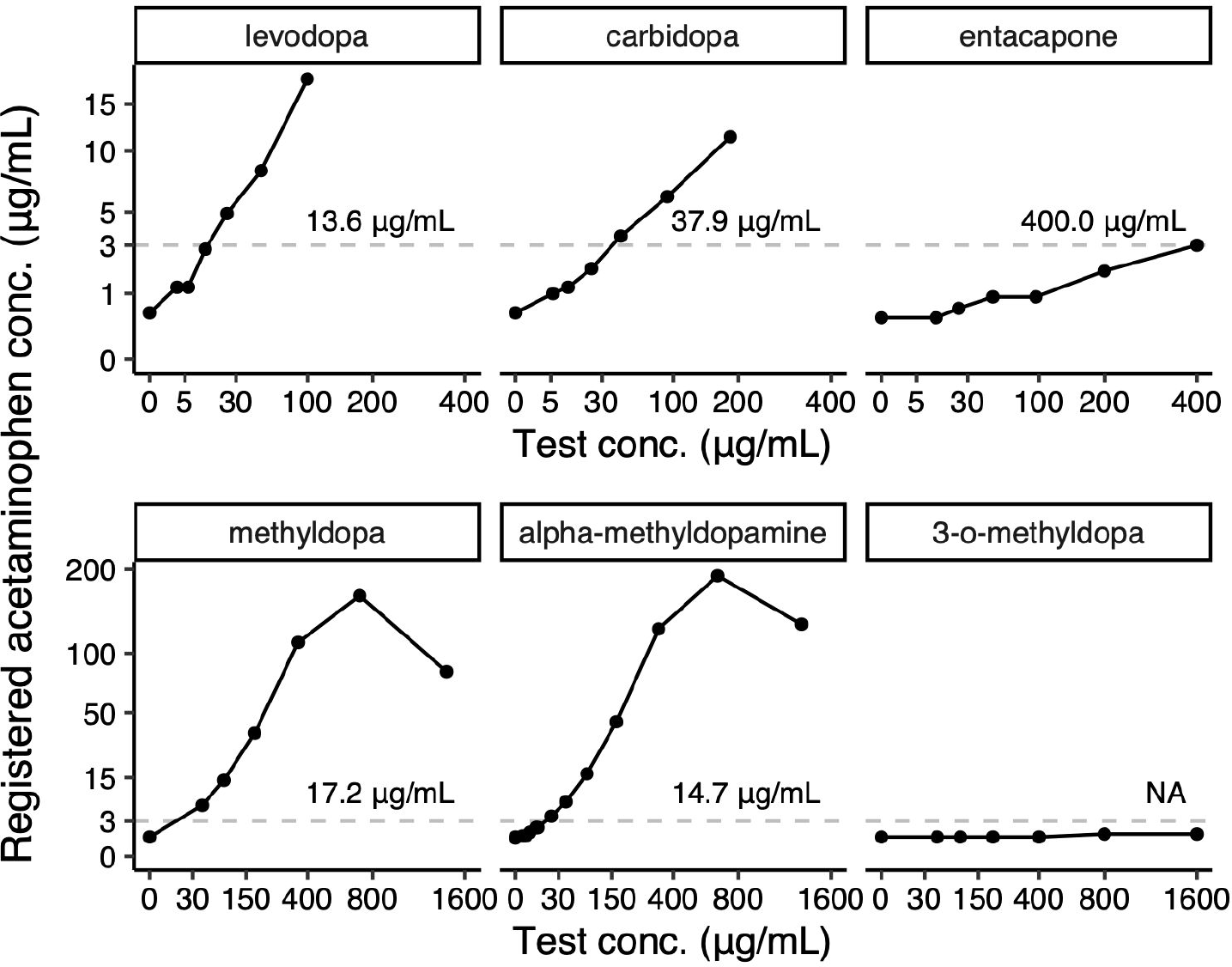
Experimental validation of interference on the acetaminophen assay. Dashed lines show the cutoff normally used to call a sample presumptive positive. The text above each dashed line indicates the estimated minimum concentration at which the test compound would produce a presumptive positive. Both axes are square-root-transformed to better show the lower concentrations.

Levodopa and carbidopa are structurally related to alpha-methyldopamine, a metabolite of methyldopa that interferes on our institution’s UDS assay for amphetamines [2]. Methyldopa was modestly associated with presumptive positive UDS results for acetaminophen (OR_P_ = 1.32, rank 44 of 854, Supplemental Table 4), and both methyldopa and alpha-methyldopamine—but not a second metabolite 3-o-methyldopa—were strongly interfering (Figure 4). Conversely, neither levodopa nor carbidopa interfered on the amphetamines assay (Supplemental Table 6). Taken together, these findings indicate that several dopamine-related compounds can cause false positive UDS results for acetaminophen.

## Discussion

Despite UDS assays’ vulnerability to interference, confirmatory testing is not always available, either for patient care or secondary analysis. Here we extended our previous approach in order to identify potentially interfering medications from EHR data without knowing which presumptive positive UDS results were true positives and which were false positives.

As EHR data are observational, our approach only quantifies associations, it does not imply causality. Thus, the decision of which compounds to experimentally evaluate should involve both the statistical analysis and clinical expertise. For example, we did not pursue the top-ranked ingredients on the ethanol assay (thiamine, diazepam, and naltrexone), because we considered it likely that the associations were due to confounding with alcohol-use disorder. In the future, accounting for such underlying patient phenotypes could further improve our approach. In any case, our findings suggest that statistical analysis of EHR data may help reveal sources of interference on a variety of clinical laboratory assays.

## Data Availability

Code and summary results for this study are available at https://doi.org/10.6084/m9.figshare.12067233.

## Acknowledgments

None declared.

## Author contributions

All the authors have accepted responsibility for the entire content of this submitted manuscript and approved submission.

## Research funding

This work was supported in part by CTSA award UL1TR002243 from NCATS/NIH. The Vanderbilt Synthetic Derivative is supported by institutional funding and by CTSA award UL1TR002243 from NCATS/NIH.

## Employment or leadership

None declared.

## Honorarium

None declared.

## Competing interests

The funding organization(s) played no role in the study design; in the collection, analysis, and interpretation of data; in the writing of the report; or in the decision to submit the report for publication

## Notes

### Competing Interest Statement

The authors have declared no competing interest.

